# Socio-Economic Factors Associated with Cancer Stigma among Apparently Healthy Women in Semi-urban Nepal

**DOI:** 10.1101/2024.03.11.24304143

**Authors:** Bandana Paneru, Aerona Karmacharya, Soniya Makaju, Diksha Kafle, Lisasha Poudel, Sushmita Mali, Priyanka Timsina, Namuna Shrestha, Dinesh Timalsena, Kalpana Chaudhary, Niroj Bhandari, Prasanna Rai, Sunila Shakya, Donna Spiegelman, Sangini S Sheth, Anne Stangl, McKenna C. Eastment, Archana Shrestha

## Abstract

Cancer is the primary cause of death globally, and despite the significant advancements in treatment and survival rates, it is still stigmatized in many parts of the world. However, there is limited public health research on cancer stigma among general population (non-patient) women in Nepal. Therefore, this study aims to determine the prevalence of cancer stigma and its associated factors in this group.

**Methods:** We conducted a cross-sectional study among 426 healthy women aged 30 – 60 years who were residents of Dhulikhel and Banepa in central Nepal. We measured cancer stigma using the Cancer Stigma Scale (CASS). CASS measures cancer stigma in six subdomains (awkwardness, avoidance, severity, personal responsibility, policy opposition, financial discrimination) on a 6-point Likert scale (strongly disagree to agree strongly) with higher mean stigma scores correlating with higher levels of stigma. We used univariable and multivariable linear regression to identify the socio-demographic factors associated with the CASS score.

**Results:** Overall, the level of cancer stigma was low (mean total stigma score: 2.6 ± 0.6) but still present among participants. Stigma related to personal responsibility had the highest levels (mean stigma score: 3.9 ± 1.3), followed by severity (mean stigma score: 3.2 ± 1.3) and financial discrimination (mean stigma score: 2.9 ± 1.6). There was a significant association of mean CASS score with older age (the mean difference is stigma score: 0.01 points; 95% CI: 0.01-0.02) and lower education (difference -0.02 points; 95% CI: -0.03, -0.003) after adjusting for age, ethnicity, education, marital status, religion, occupation, and parity.

**Conclusion:** While overall cancer stigma was low in Nepal, some subdomains were increased in the general population of women in Nepal. Because stigma may impact engagement in cancer screening efforts, programs should aim to counteract stigma, particularly among older and less educated women.

## INTRODUCTION

Cancer is the leading cause of death worldwide, with age-standardized mortality of 100 per 100,000 population in 2020.^1^ The majority of cancer deaths occur in low- and middle-income countries.^1^ In Nepal in 2020, the age-standardized cancer incidence rate was 80 per 100,000 with a mortality rate of 54 per 100,000 in 2020.^1^ Despite recent improvements in treatment and survival, cancer is still a stigmatized disease,^2–4^ and one of the most feared illnesses.^5^

Health-related stigma subjects a person or group to exclusion, rejection, blame, or devaluation due to the anticipation or experience of negative social judgment regarding their health condition, making it a social phenomenon or personal experience.^6^ Public stigma may appear in the form of stereotypes, for instance, viewing individuals who have survived cancer as either incapable or contagious. These stereotypes result in behaviors like avoiding interactions with cancer survivors due to a fear of contracting the disease. Discriminatory actions stemming from these biases can lead to withholding job opportunities or rejecting social interactions.^7–9^ This public stigma significantly hinders individuals’ willingness to seek health care.^6,10^

Prior research has predominantly examined stigma related to illnesses such as leprosy,^11,12^ epilepsy, HIV/AIDS,^13,14^ and mental illness.^15–17^ Although people often stigmatize cancer, there is a limited exploration of public stigma in healthy general non-patient populations.^10^ Examining public stigma related to cancer is crucial for several reasons. First, stigma could dissuade individuals from participating in cancer prevention and screening efforts, resulting in delayed cancer diagnoses and, ultimately, higher mortality rates.^10,18–20^ Second, public health initiatives aimed at educating people about the behavioral factors linked to cancer, including smoking, obesity, and infection with the human papillomavirus, could create stigma by suggesting that cancer is avoidable and depends on individual behavior and choices.^21^ Third, stigma can contribute to health disparities,^22–24^ particularly among marginalized groups who may already face barriers to accessing healthcare.

Few studies have actively investigated cancer-related stigma and associated socio-demographic factors. Qualitative research delving into experienced and internalized stigma among cancer patients reveals that young, single individuals encounter distinct stigma experiences influenced by their age, gender, marital status, socio-economic position, and family living arrangements.^25,26^ A quantitative exploration of stigma in England revealed a higher prevalence of cancer stigma among men and individuals from ethnic minority backgrounds but no associations with age or social status. Notably, these studies utilized only 18 of the 25 CASS available items, potentially resulting in low internal reliability. Moreover, studies have yet to be conducted in low-income countries, notably Nepal. This study aimed to determine the prevalence of cancer stigma and its associated factors among women residing in semi-urban parts of Nepal. By identifying these factors, we can develop interventions and policies that can help reduce stigma and its adverse effects on individuals and society and ultimately improve the population’s health, where cancer screening and early treatment are lifesaving.

## METHODS

### Study design and setting

We conducted a cross-sectional study in two municipalities of the Kavrepalanchow district of Nepal, Dhulikhel and Banepa, approximately 30 kilometers east of Kathmandu. Dhulikhel is a semi-urban location with a population of 32,162, while Banepa has a population of 55,628. The dominant ethnic group in both municipalities is the Newar, and 70% of the population is literate. The literacy rate among females is around 75% in both municipalities.^27^

### Study Participants

Our study population included women aged 30 to 60 years who were residents of Dhulikhel or Banepa. The exclusion criteria were: a) having a hearing impairment, b) having severe mental health conditions so as being not able to provide informed consent, and c) not being a resident of Dhulikhel or Banepa (i.e., visitors to the area).

### Recruitment

We enrolled the initial 426 women out of 1800 who underwent cervical cancer screening organized by Dhulikhel Hospital from May 15 to September 15, 2021. We estimated the sample size based on an expected proportion of cancer stigma among women of 51%,^10^ at a 5% significance level, and a margin of error of 5%.^28^

Our Research Assistants (RAs) contacted female community health volunteers (FCHV) and oriented them to the study objective and expectations. These FCHVs have worked in Nepal since the 1980s and play a pivotal role in the Nepali community health workforce, specializing in health education, counseling, outreach, and resource distribution.^29^ The FCHVs disseminated information about the study to women in their network and shared the contact details of the interested participants with the research assistants. Subsequently, the research assistants contacted the women by phone, outlining the study’s objectives and explaining their potential role. Women expressing interest were then formally enrolled in the study after verbal informed consent was obtained, ensuring the anonymity and confidentiality of their information. We conducted the study amid the COVID-19 pandemic. Therefore, interviews were conducted over the phone for infection prevention. Kathmandu University Institutional Review Committee ethical board approved the study (KUIRC no: 35/2021).

### Data collection

Trained research assistants conducted telephonic interviews using a structured questionnaire directly entered electronically (Kobo toolbox).^30^ The questionnaire covered socio-demographic factors and cancer stigma.

### Measures

#### Cancer Stigma

We used Nepal’s validated Cancer Stigma Scale (CASS).^31^ The CASS demonstrated satisfactory internal reliability, with a Cronbach’s alpha of 0.88 for the overall scale and ranging from 0.70 to 0.89 for its six components.^31^ The CASS consists of 25 items that assess six domains: (a) awkwardness, which measures how comfortable people feel around someone with cancer; (b) severity, which evaluates the expected severity of cancer consequences and the likelihood of recovery; (c) avoidance, which assesses the extent to which people avoid cancer patients and maintain physical distance from them; (d) personal responsibility, which determines how much a person’s actions contribute to their cancer; (e) policy opposition, which gauges the perceived responsibility of the government and the public in the care and treatment of cancer patients; and (f) financial discrimination, which measures the anticipated deprivation of benefits to cancer patients from banking and insurance services

Participants responded using a 6-point Likert scale ranging from ‘disagree strongly’ to ‘agree strongly.’ We reversed the scores for five specific items, ensuring higher scores reflected more stigma levels (refer to Table 2 for details on the reverse-scored items).^10,17^ The mean score for each domain was then calculated.^10,17^

**Table 1:** Socio-demographic characteristics of the study participants (n=426)

| Characteristics | Frequency (%) |
| --- | --- |
| Age (in years), Mean(SD) | 42.4 (8.2) |
| Ethnicity |  |
| Brahmin/Chettri/Thakuri/Sanyasi | 182 (42.7) |
| Newar | 175 (41.1) |
| Magar/Tamang/Rai/Limbu | 13 (3.1) |
| Sherpa/Bhote | 27 (6.3) |
| Kami/Damai/Sarki/Gaaine/Baadi | 24 (5.6) |
| Others | 5 (1.2) |
| Religion |  |
| Hindu | 374 (87.8) |
| Buddhist | 28 (6.6) |
| Christian | 24 (5.6) |
| Education |  |
| No formal education | 132 (31.0) |
| Primary | 49 (11.5) |
| Secondary | 150 (35.2) |
| Above secondary | 95 (22.3) |
| Occupation |  |
| Farmer | 168 (39.5) |
| Home-maker | 106 (24.9) |
| Business | 63 (14.8) |
| Unemployed | 7 (1.6) |
| Other | 82 (19.2) |
| Parity (number), Mean (SD) | 2.3 (1.0) |

**Table 2.** Cancer stigma scale (CASS) mean score in each domain among participants (n=426)

|  | Mean(sd) |
| --- | --- |
| **<u>Awkwardness</u>** | **2.4 (1.2)** |
| I would feel at ease around someone with cancer (R) | 2.4 (1.7) |
| I would feel comfortable around someone with cancer (R) | 2.4 (1.7) |
| I would find it difficult being around someone with cancer | 2.3 (1.7) |
| I would find it hard to talk to someone with cancer | 2.2 (1.6) |
| I would feel embarrassed discussing cancer with someone who had it | 2.7 (1.8) |
| **<u>Severity</u>** | **3.2 (1.3)** |
| Once you've had cancer, you're never 'normal' again | 3.2 (1.7) |
| Having cancer usually ruins a person's career | 3.4 (1.7) |
| Getting cancer means having to mentally prepare oneself for death | 3.4 (1.7) |
| Cancer usually ruins close personal relationships | 3.1 (1.7) |
| Cancer devastates the lives of those it touches | 2.8 (1.7) |
| **<u>Avoidance</u>** | **1.7 (0.9)** |
| If a colleague had cancer, I would try to avoid them | 1.7 (1.2) |
| I would distance myself physically from someone with cancer | 1.8 (1.4) |
| I would feel irritated by someone with cancer | 1.3 (0.8) |
| I would feel angered by someone with cancer | 1.2 (0.7) |
| I would try to avoid a person with cancer | 2.1 (1.7) |
| **<u>Policy Opposition</u>** | **1.3 (0.6)** |
| More government funding should be spent on the care and treatment of those with cancer (R) | 1.3 (0.8) |
| The needs of people with cancer should be given top priority (R) | 1.2 (0.5) |
| We have a responsibility to provide the best possible care for people with cancer (R) | 1.3 (0.6) |
| **<u>Personal Responsibility</u>** | **3.9 (1.3)** |
| A person with cancer is liable for their condition | 4.2 (1.6) |
| A person with cancer is accountable for their condition | 4.4 (1.5) |
| If a person has cancer, it's probably their fault | 3.6 (1.6) |
| A person with cancer is to blame for their condition | 3.2 (1.6) |
| **<u>Financial discrimination</u>** | **2.9 (1.6)** |
| It is acceptable for banks to refuse to make loans to people with cancer | 2.4 (1.8) |
| Banks should be allowed to refuse mortgage applications for cancer-related reasons | 2.6 (1.8) |
| It is acceptable for insurance companies to reconsider a policy if someone has cancer | 3.7 (1.9) |
| **<u>Overall stigma</u>** | **2.6 (0.6)** |

\*Stigma score ranges from 1-6. Higher scores indicate a higher sigma level. (R) indicated reversed in scoring.

**Table 3:** Factors associated with cancer stigma score among participants (n=426)

| Characteristics | Univariable<br>Difference in CASS<br>score (95% CI) | p-value | Multivariable<br>Difference in CASS score<br>(95% CI) | p-value |
| --- | --- | --- | --- | --- |
| *Age (years), Mean (SD) | 0.2 (0.1, 0.3) | 0.002 | 0.18 (0.1, 0.2) | 0.02 |
| Education (years of formal<br>education), Mean (SD) | 0.04 (-0.05, -0.02) | <0.001 | -0.02 (-0.03, -0.01) | 0.02 |
| Ethnicity |  |  |  |  |
| Brahmin/Chhetri | Ref | 0.67 |  | 0.54 |
| Damai/Sarki/Gaaine/Baadi | 0.03 (-0.39, 0.40) |  | -0.12 (-0.42, 0.17) |  |
| Magar/Tamang/Rai/Limbu | 0.19 (-0.70, 0.37) |  | -0.27 (- 0.56, 0.02) |  |
| Newar | -0.08 (-0.29, 0.12) |  | 0.04 (-0.10, 0.19) |  |
| Sherpa/Bhote | 0.09 (-0.31, 0.50) |  | 0.03 (-0.33, 0.41) |  |
| Others | 0.78 (-0.12, 1.69) |  | 0.03 (- 0.49, 0.56) |  |
| Occupation |  |  |  |  |
| Business | Ref | 0.53 |  | 0.93 |
| Farmer | 0.37(0.08, 0.67) |  | -0.01 (-0.19, 0.18) |  |
| Home-maker | 0.02 (-0.29, 0.33) |  | -0.07 (-0.26, 0.11) |  |
| Others | 0.15 (-0.18, 0.48) |  | 0.03 (-0.17, 0.23) |  |
| Unemployed | 0.59 (-0.20, 1.38) |  | 0.07 (-0.27, 0.41) |  |
| Parity, Mean (SD) | 0.09 (-0.01, 0.18) |  | -0.01 ( -0.08, 0.07) |  |
| Religion |  |  |  |  |
| Buddhist | Ref | 0.04 |  | 0.15 |
| Christian | -0.02 (-0.57, 0.63) |  | -0.14 (-0.63, 0.35) |  |
| Hindu | -0.19 (-0.59, 0.19) |  | -0.20 (-0.50, 0.09) |  |

CI: Confidence interval
Adjusting variables- age, education, ethnicity, occupation, parity and religion
\*Age coefficients presented in 10 years difference

#### Socio-demographic variables

Socio-demographic variables included age (in years), ethnicity (Brahmin/ Chettri/Thakuri/Sanyasi, Newar, Magar/Tamang/Rai/Limbu, Sherpa/Bhote, Kami/Damai/Sarki/Gaaine/Baadi, Other), education (number of years of formal education completed), religion (Hindu, Buddhist, Christian), occupation (homemaker, farmer, business, unemployed, others) and parity (number of children). We adopted the questions from previously conducted national surveys in Nepal. Responses were self-reported.^32,33^

### Data analysis

We calculated summary statistics such as frequency and percentage for categorical variables and mean (standard deviation) for continuous variables. We estimated cancer stigma scores for six sub-domains and calculated mean scores for each subscale. We performed multivariate linear regression analysis to determine the socio-demographic factors associated with cancer stigma, selecting the variables in our model based on literature review^10,20^ and prior knowledge. We reported crude and adjusted differences in stigma scores and their 95% confidence intervals and p-values. We conducted all analyses using STATA-13.

## RESULTS

Table 1 presents the socio-demographic characteristics of the participants. The mean age was 42 ± 8 years. Most participants (43%) were Brahmin/Chhetri and Hindus (87.8%). About a third of the participants (31%) had no formal education, and the majority (39.5%) were farmers. The mean number of children was 2.3 (SD 1.0).

Table 2 presents the cancer stigma scale (CASS) mean score for each stigma domain. The overall mean total CASS score was 2.6 ± 0.6. Within the six domains, the highest stigma level pertained to personal responsibility, reflecting the belief that patients are accountable for acquiring cancer (mean stigma score: 3.9 ± 1.3). Additionally, high stigma levels were observed in the severity domain, indicating that patients may struggle to return to everyday life, adversely affecting their overall life and relationships (mean stigma score: 3.2 ± 1.3). Financial discrimination, manifested by the denial of loans and mortgage applications for cancer patients, also exhibited a notable stigma level (mean stigma score: 2.9 ± 1.6). Policy opposition showed a low mean stigma score (1.3 ± 0.6), suggesting strong government and community support for cancer patient care.

In the univariable regression model, the mean cancer stigma score showed associations with age, with older women having a higher mean stigma score, education, with more highly educated women having a lower mean stigma score, and occupation, with farmers having higher mean stigma scores compared to businesswomen. However, after adjusting for socio-demographic variables, the mean cancer stigma score was only associated with age and education. When comparing two groups of individuals differing by ten years of age, the older group had a 0.18 unit higher cancer stigma score compared to the younger group after adjusting for education, ethnicity, occupation, parity, and religion (95% confidence interval [CI]: 0.04-0.20; p-value=0.04). There was a significant negative association between education and the mean cancer stigma score (p-value=0.03). The mean cancer stigma score was 0.037 units lower with one year more formal education among women (95% CI: -0.032--0.001). Cancer stigma score was not significantly associated with other socio-demographic variables such as ethnicity, occupation, parity, and religion (p>0.05).

## DISCUSSION

Our study found a low level of general cancer stigma that exists among apparently healthy women in semi-urban Nepal. We observed higher levels of cancer stigma for the domains of personal responsibility, severity, and financial discrimination. Regarding personal responsibility, participants believed that cancer patients bore responsibility for the onset of their illness.

Likewise, in the severity domain, women conveyed perceptions of cancer as a terminal ailment, with individuals never regaining an everyday life. Furthermore, the financial discrimination domains underscored participants’ endorsement of the idea that banks could justifiably refuse loans and mortgages based on cancer-related reasons. Cancer stigma scores were higher among older individuals and those with lower levels of formal education.

In previous studies exploring cancer stigma, consistently low levels of stigma have been reported.^10,35^ A cross-sectional study conducted in the UK reported an overall mean cancer stigma score higher than the one observed in our study.^10^ In contrast to women from the UK, our study population exhibited a higher total mean cancer stigma score across five sub-domains: severity, awkwardness, financial discrimination, personal responsibility, and avoidance.

Conversely, our study participants demonstrated a lower mean stigma score in the sub-domain of policy opposition than English women, indicating less support for government funding toward cancer care and treatment.^10,17^ Stigma varies across diseases, even among different types of cancers. HIV patients encounter more significant stigma than cancer patients, leading to more reported psychological dysfunctions.^34^ In a study of 1205 non-patient participants in the UK using the CASS tool, lung cancer showed higher stigma scores than breast, cervical, skin, and colorectal cancers. ^35^

The low mean cancer stigma score observed in the policy opposition sub-domain suggests a distinct perspective in our research. Unlike the findings from the English study,^10^ Nepali participants did not anticipate receiving substantial support from the government or community for cancer diagnosis and treatment. This difference can be attributed to Nepal and England’s different health financing mechanisms and community structures. In Nepal, out-of-pocket healthcare expenditure is notably high (55%),^36^ compared to England (12.5%).^37^ Our study participants may have experienced a greater need for government support for cancer treatment. Moreover, Nepal’s more cohesive community structure might have inclined our participants to prioritize the shared responsibility of caring for cancer patients within their community. These findings emphasize the significance of contextual factors in shaping attitudes and perceptions related to cancer stigma across different settings.

The study revealed a significant inverse association between education and cancer stigma among participants. Similar findings were reported in studies conducted in Ireland^38^ and China^39^ where individuals with lower levels of education had higher cancer stigma scores. These findings imply that the link between education and cancer stigma may exist across different cultural contexts and regions. Those with lower levels of education may have limited access to health literacy and information on health-related matters, potentially leading to misconceptions and misinformation.^55^

Our findings suggest a positive association between older age and cancer stigma among women in Nepal, which is in contrast to studies conducted in England,^10^ China,^40^ and Kenya.^20^ This difference might be explained by the comparatively lower exposure to social media platforms. Specifically, 2.2% of Nepali women aged 50 and above use social media.^41^ In the UK, media exposure is considerably higher, with 50% of individuals using social media at 50,^42^ while in China, 41.5% of social media users are aged above 40 years.^43^ Social media might reduce stigma, fostering awareness, empathy, and community support by disseminating accurate information, personal stories, and advocacy efforts.^44^ This highlights the need for targeted educational interventions to improve knowledge and awareness about cancer among older women in Nepal.

This is the first quantitative study conducted in Nepal among healthy adult women to identify the factors associated with cancer stigma. A previous qualitative study in Nepal focused on a limited pool of people with cancer, exploring only a few dimensions of stigma.^45^ The current research’s strength lies in applying a quantitative approach that introduces objectivity to measure diverse stigma domains among healthy women who have not experienced cancer themselves. In addition, we used CASS, a standardized tool validated in Nepal, to collect our data to estimate the factors associated with cancer stigma.^31^ We utilized multivariate linear regression models, adjusting for potential confounders, including age, education, ethnicity, occupation, parity, and religion, which helps to eliminate alternative explanations of our findings.

This study has a few limitations. First, response bias is possible due to the interviewer-administered nature of the surveys. Respondents might have downplayed negative emotions in their responses, possibly causing an underreporting of their experiences and, consequently, our stigma score.^46^ Second, convenience sampling techniques may introduce selection bias, limiting our findings’ generalizability to the Nepalese female population. Future research with a random sample of women is needed to confirm our findings. Third, the study’s cross-sectional design provides a snapshot of cancer stigma at a specific time. It cannot determine changes in stigma over time. Fourth, this study employed CASS items to evaluate cancer stigma in general. Nevertheless, stigma may vary across different cancer types.^35^ Consequently, future research should strive to explore the stigma associated with specific cancer types and their determinants.

In conclusion, this study revealed that the overall cancer stigma was low but still exists among women in a suburban area in central Nepal. Stigma may impact engagement in cancer screening efforts, so stigma reduction intervention focusing an older and less educated women is recommended.

## Data Availability

Data are available within the manuscript as a supporting Information files

## Supporting Information

**S1 Dataset**

**S2 Data collection tool**

